# Inferring the effective start dates of non-pharmaceutical interventions during COVID-19 outbreaks

**DOI:** 10.1101/2020.05.24.20092817

**Authors:** Ilia Kohanovski, Uri Obolski, Yoav Ram

## Abstract

During Feb-Apr 2020, many countries implemented non-pharmaceutical interventions, such as school closures and lockdowns, with variable schedules, to control the COVID-19 pandemic caused by the SARS-CoV-2 virus. Overall, these interventions seem to have successfully reduced the spread of the pandemic. We hypothesise that the official and effective start date of such interventions can significantly differ, for example due to slow adoption by the population, or because the authorities and the public are unprepared. We fit an SEIR model to case data from 12 countries to infer the effective start dates of interventions and contrast them with the official dates. We find mostly late, but also early effects of interventions. For example, Italy implemented a nationwide lockdown on Mar 11, but we infer the effective date on Mar 17 (±2.99 days 95% CI). In contrast, Germany announced a lockdown on Mar 22, but we infer an effective start date on Mar 19 (± 1.05 days 95% CI). We demonstrate that differences between the official and effective start of NPIs can distort conclusions about their impact, and discuss potential causes and consequences of our results.

## Introduction

The COVID-19 pandemic has resulted in implementation of extreme non-pharmaceutical interventions (NPIs) in many affected countries. These interventions, from social distancing to lockdowns, are applied in a rapid and widespread fashion. NPIs are designed and assessed using epidemiological models, which follow the dynamics of infection to forecast the effect of different mitigation and suppression strategies on the levels of infection, hospitalization, and fatality. These epidemiological models usually assume that the effect of NPIs on infection dynamics begins at the officially declared date^8,10,15^.

Adoption of public-health recommendations is often critical for effective response to infectious diseases, and has been studied in the context of HIV^14^ and vaccination^5,20^, for example. However, behavioural and social change does not occur immediately, but rather requires time to diffuse in the population through media, social networks, and social interactions. Moreover, compliance to NPIs may differ between different interventions and between people with different backgrounds. For example, in a survey of 2,108 adults in the UK during Mar 2020, Atchison et al.^2^ found that those over 70 years old were more likely to adopt social distancing than young adults (18-34 years old), and that those with lower income were less likely to be able to work from home and to self-isolate. Similarly, compliance to NPIs may be impacted by personal experiences. Smith et al.^17^ have surveyed 6,149 UK adults in late Apr 2020 and found that people who believe they have already had COVID-19 are more likely to think they are immune, and less likely to comply with social distancing guidelines. Compliance may also depend on risk perception as perceived by the the number of domestic cases or even by reported cases in other regions and countries. Interestingly, the perceived risk of COVID-19 infection has likely caused a reduction in the number of influenza-like illness cases in the US starting from mid-Feb 2020^21^.

Here, we hypothesise that there is a significant difference between the official start of NPIs and their effective adoption by the public and therefore their effect on infection dynamics. We use a *Susceptible-Exposed-Infected-Recovered* (SEIR) model and a *Markov Chain Monte Carlo* (MCMC) parameter estimation framework to infer the effective start date of NPIs from publicly available COVID-19 case data in 12 geographical regions. We compare these estimates to the official dates, and find that they include both late and early effects of NPIs on infection dynamics. We conclude by demonstrating how differences between the official and effective start of NPIs can confound assessments of their impacts.

## Results

Several studies have described the effects of non-pharmaceutical interventions in different geographical regions^8,10,15^. Some of these studies have assumed that the parameters of the epidemiological model change at a specific date (Eq. 6), and set the change date *τ* to the official NPI date *τ*∗ (Table 1). They then fit the model once for time *t* < *τ*∗ and once for time *t* ≥ *τ*∗. For example, Li et al.^15^ estimate the infection dynamics in China before and after *τ*∗, which is set at Jan 23, 2020. Thereby, they effectively estimate the transmission and reporting rates before and after *τ*∗ separately.

**Table 1:** Official start of non-pharmaceutical interventions. The date of the first intervention is for a ban of public events, or encouragement of social distancing, or for school closures. In all countries except Sweden, the date of the last intervention is for a lockdown. In Sweden, where a lockdown was not ordered during the studied dates, the last date is for school closures. Dates for European countries from Flaxman et al.^8^, date for Wuhan, China from Pei and Shaman^16^. See Figure S1 for a visual presentation.

| Country | First | Last |
| --- | --- | --- |
| Austria | Mar 10 2020 | Mar 16 2020 |
| Belgium | Mar 12 2020 | Mar 18 2020 |
| Denmark | Mar 12 2020 | Mar 18 2020 |
| France | Mar 13 2020 | Mar 17 2020 |
| Germany | Mar 12 2020 | Mar 22 2020 |
| Italy | Mar 5 2020 | Mar 11 2020 |
| Norway | Mar 12 2020 | Mar 24 2020 |
| Spain | Mar 9 2020 | Mar 14 2020 |
| Sweden | Mar 12 2020 | Mar 18 2020 |
| Switzerland | Mar 13 2020 | Mar 20 2020 |
| United Kingdom | Mar 16 2020 | Mar 24 2020 |
| Wuhan | Jan 23 2020 | Jan 23 2020 |

Here, we estimate the joint posterior distribution of the effective start date of NPIs, *τ*, and the transmission and reporting rates before and after *τ* from the entire data, rather than splitting the data at *τ*. We then estimate 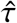 as the median of the marginal posterior distribution of *τ*. This is done under the simplifying assumption that all interventions start at a specific date, despite the reality that the durations between the first and last NPIs were between 4 and 12 days (Table 1, Figure S1).

We compare the posterior predictive plots of a model with a free *τ* with those of a model with *τ* fixed at *τ*∗ and a model without *τ* (i.e. transmission and reporting rates are constant). All three models were fitted to case data up to Apr 11, used to predict out-of-sample case data up to Apr 24, and these predictions were then compared to the real case data. The model with free *τ* clearly produces better and less variable predictions (Figure S4a): In all 11 of the European countries, the expected posterior RMSE (root mean squared error) of the out-of-sample predictions is lowest for the model with a free *τ* (Table S2). When we compare the models using WAIC (Eq. 10), the model with a free *τ* is strongly preferred in 9 out of 12 countries, the exceptions being Norway (only slight preference), Sweden, and Denmark (Table S1). Notably, the data for Sweden and Denmark do not have a single “peak” during the evaluated dates, possibly leading to wide 95% credible intervals on 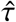 (Figure 1) and poor WAIC in the model with free *τ*, whereas the duration between the first and last interventions was especially long in Norway (Table 1).

**Figure 1:**
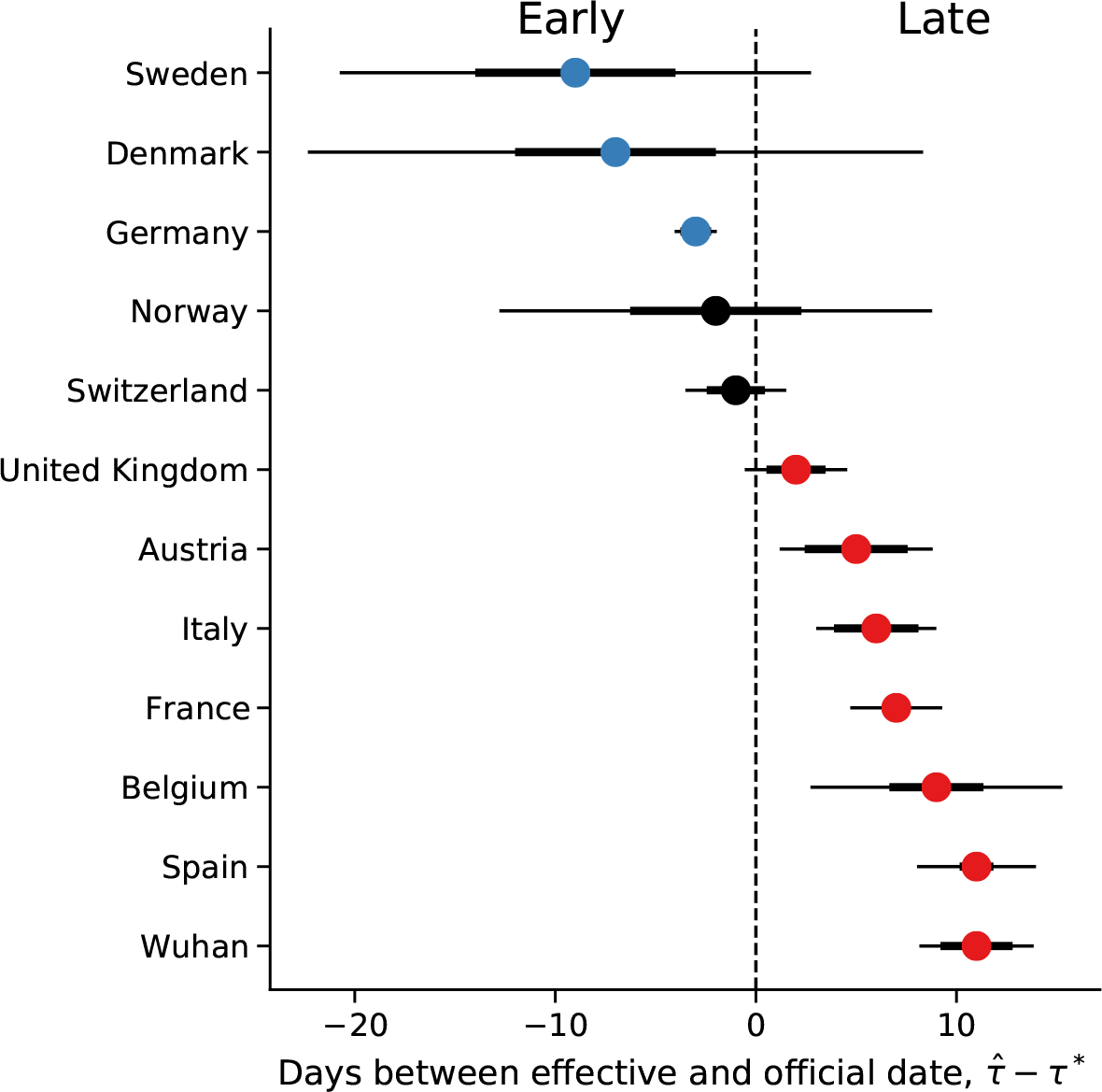
Official vs. effective start of non-pharmaceutical interventions. The difference between 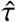 the effective and *τ*∗ the official start of NPIs is shown for different regions. The effective date is delayed in UK, Austria, Italy, France, Belgium, Spain, and Wuhan, China, compared to the official date (red markers). In contrast, the estimated effective dates in Sweden, Denmark, and Germany are earlier than the official dates (blue markers), although uncertainty is low only for Germany (i.e., zero is not included in 95% CI). The credible intervals for Sweden, Denmark, and Norway are especially wide, see text and Figure 3 for possible explanation. Here, 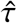 is the posterior median, see Table 2. *τ*∗ is the last NPI date (Table 1). Thin and bold lines show 95% and 75% credible intervals, respectively. Figure S2 shows a similar summary when estimating 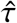 using case data up to Mar 28, 2020, rather then Apr 11.

We compare the official *τ*∗ and effective 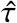 start of NPIs and find that in most regions the effective start of NPIs differs from the official date: in 10 of 12 countries the 75% credible interval on 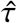 does not include *τ*∗ (7 of 12 countries when considering a 95% interval; Figure 1). The exceptions are Switzerland and Norway (see below). The latter also has a wide 95% credible interval, perhaps because it has the longest duration between the first and last NPIs (Table 1).

**Table 2:** Parameter estimates for different regions. See Eq. 1 for model parameters. All estimates are posterior medians. 75% and 95% credible intervals given for *τ*, in days. *τ*∗ is the official last NPI date, see Table 1.

| Country | $\tau^*$ | $\tau$ | $CI_{75\%}$ | $CI_{95\%}$ | Z | D | $\mu$ | $\beta$ | $\alpha_1$ | $\lambda$ | $\alpha_2$ | E(0) | $I_u(0)$ | $\Delta t$ |
| --- | --- | --- | --- | --- | --- | --- | --- | --- | --- | --- | --- | --- | --- | --- |
| Austria | Mar 16 | Mar 21 | 2.5658 | 3.8148 | 3.9102 | 3.6649 | 0.6407 | 1.1417 | 0.1308 | 0.1387 | 0.2261 | 151.1078 | 131.6623 | 2.1308 |
| Belgium | Mar 18 | Mar 27 | 2.3456 | 6.2773 | 3.9098 | 3.6152 | 0.4877 | 1.0762 | 0.2721 | 0.4388 | 0.2363 | 309.9114 | 426.9644 | 2.1756 |
| Denmark | Mar 18 | Mar 11 | 5.0053 | 15.3374 | 4.0957 | 3.5091 | 0.4161 | 1.0747 | 0.1470 | 0.6919 | 0.1867 | 324.6332 | 389.5076 | 2.2071 |
| France | Mar 17 | Mar 24 | 0.5068 | 2.2928 | 4.2159 | 3.1688 | 0.4863 | 1.0502 | 0.3726 | 0.3765 | 0.4168 | 422.6720 | 1362.5052 | 1.6717 |
| Germany | Mar 22 | Mar 19 | 0.7657 | 1.0508 | 3.3917 | 3.7166 | 0.6780 | 1.1515 | 0.1753 | 0.2664 | 0.4147 | 529.9397 | 387.1038 | 2.1516 |
| Italy | Mar 11 | Mar 17 | 2.1045 | 2.9945 | 4.1439 | 2.5550 | 0.5371 | 0.9932 | 0.5816 | 0.3860 | 0.4973 | 990.1942 | 1902.7806 | 1.6367 |
| Norway | Mar 24 | Mar 22 | 4.2700 | 10.7889 | 4.0476 | 3.1061 | 0.3885 | 1.0467 | 0.1536 | 0.3091 | 0.2320 | 478.0678 | 856.7700 | 1.9998 |
| Spain | Mar 14 | Mar 25 | 0.8415 | 2.9656 | 4.1392 | 3.3352 | 0.5974 | 1.1574 | 0.3640 | 0.2551 | 0.3050 | 310.0470 | 957.8280 | 1.6062 |
| Sweden | Mar 18 | Mar 09 | 4.9930 | 11.7493 | 4.0328 | 3.3962 | 0.3656 | 1.0457 | 0.1427 | 0.7237 | 0.3093 | 394.0202 | 543.5448 | 2.3755 |
| Switzerland | Mar 20 | Mar 19 | 1.4513 | 2.5140 | 3.9764 | 3.4425 | 0.6360 | 1.1543 | 0.1283 | 0.2481 | 0.2246 | 336.3842 | 328.7800 | 1.9891 |
| United Kingdom | Mar 24 | Mar 26 | 1.4721 | 2.5543 | 3.9654 | 3.6333 | 0.6414 | 1.1168 | 0.1212 | 0.4504 | 0.1737 | 422.8841 | 486.0467 | 2.0540 |
| Wuhan, China | Jan 23 | Feb 03 | 1.7984 | 2.8493 | 3.7326 | 3.6320 | 0.6057 | 1.1453 | 0.2754 | 0.1784 | 0.3511 | 597.8676 | 561.1586 | 2.4248 |

In the following, we describe our findings in more detail.

### Late effective start of NPIs

In half of the examined regions, we estimate that the effective start of NPIs 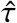 is later than the official date *τ*∗.

In Italy, the first case was officially confirmed on Feb 21. School closures were implemented on Mar 5^8^, a lockdown was declared in Northern Italy on Mar 8, with social distancing implemented in the rest of the country, and the lockdown was extended to the entire nation on Mar 11^10^. That is, the first and last official NPI dates are Mar 5 and Mar 11. However, we estimate the effective date 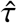 six days after the lockdown, at Mar 17 (±2.99 days 95% CI; Figure 2).

**Figure 2:**
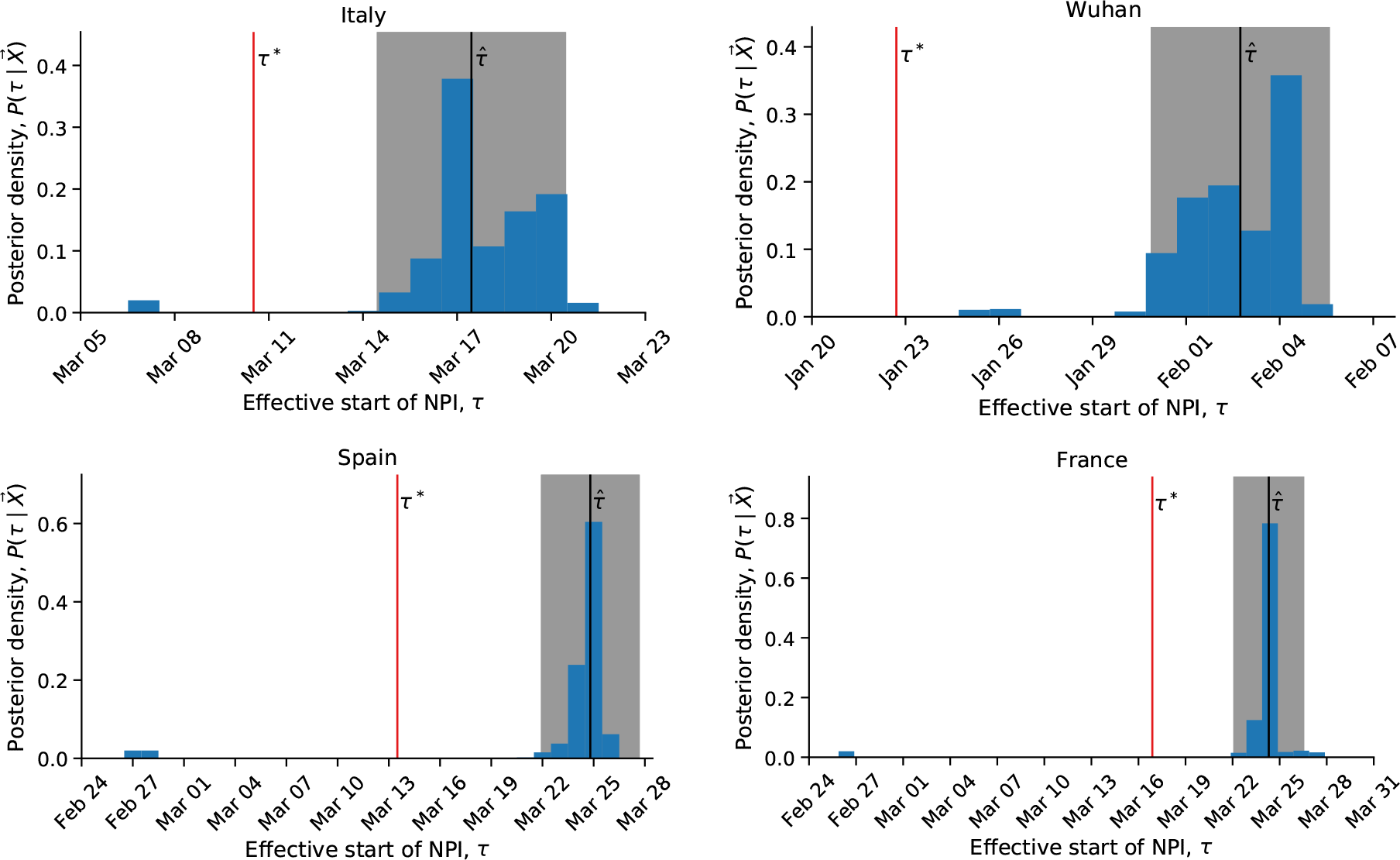
Late effect of non-pharmaceutical interventions. Posterior distribution of *τ*, the effective start date of NPI, is shown as a histogram of MCMC samples. Red line shows the official last NPI date *τ*∗. Black line shows the estimate 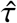. Shaded area shows a 95% credible interval.

In Wuhan, China, a lockdown was ordered on Jan 23^15^, but we estimate the effective start of NPIs to be more than a week later, at Feb 3 (±2.85 days 95% CI; Figure 2).

In Spain, social distancing was encouraged starting on Mar 8^8^, but mass gatherings still occurred on Mar 8, including a march of 120,000 people for the International Women’s Day, and a football match between Real Betis and Real Madrid (final score 2–1) with a crowd of 50,965 in Seville. A national lockdown was only announced on Mar 14^8^. Nevertheless, we estimate the effective start of NPI 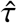 ten days later, at Mar 24 (±2.96 days 95 %CI), rather than Mar 14 (Figure 2).

Similarly, in France we estimate the effective start of NPIs 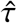 at Mar 24 (± 2.29 days 95% CI, Figure 2). This is a week later than the official lockdown, which started at Mar 17, and more than 10 days after the earliest NPI, banning of public events, which started on Mar 13^8^.

Interestingly, the effective start of of NPIs 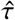 in both France and Spain is estimated to have started on Mar 24, although the official NPI dates differ significantly: the first NPI in France is only one day before the last NPI in Spain. The number of daily cases was similar in both countries until Mar 8, but diverged by Mar 13, reaching much higher numbers in Spain (Figure S3). This may suggest correlations between effective starts of NPIs due to global or international events.

### Early effective start of NPIs

In some regions we estimate an effective start of NPIs 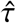 that is *earlier* then the official date *τ*∗ (Figure 1). The only conclusive early case is Germany, in which we estimate the effective start of NPIs at Mar 19 (± 1.05 days 95 %CI; Figure 3). This estimate falls between the first and last official NPI dates, Mar 12 and Mar 22 (Table 1). Therefore, when we refer to this case as “early”, we mean that the effective date (Mar 19) occurs *before* the official lockdown date (Mar 22), not that it occurs before all the NPIs. Interestingly, Germany has the second longest duration between first and last NPIs after Norway (10 and 12 days respectively; see Table 1). Nevertheless, the credible interval for the effective date in Germany is narrow (±1.05 days 95 %CI), whereas it is quite wide in Norway (±10.79 days; Figure 3).

**Figure 3:**
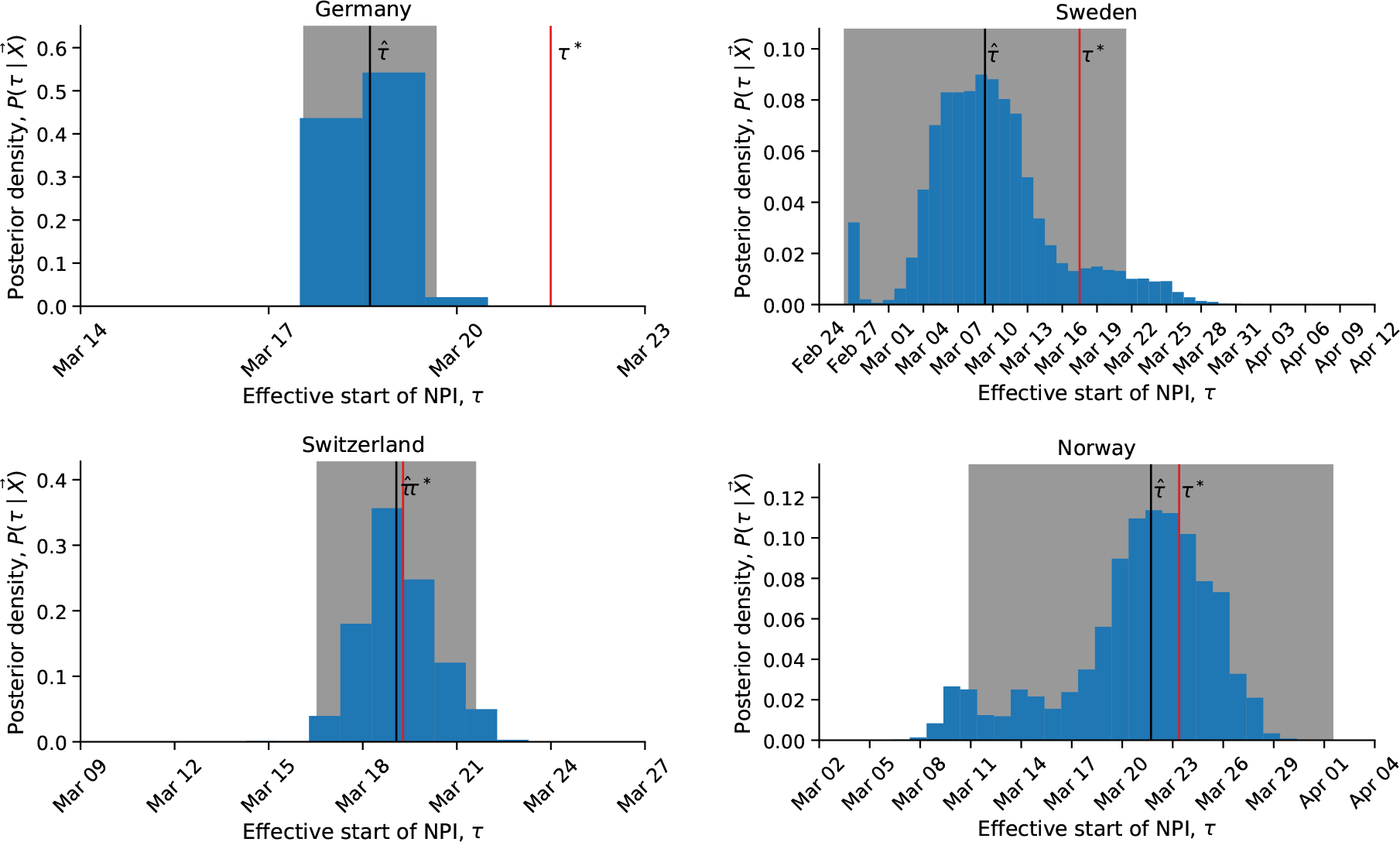
Early and exact effect of non-pharmaceutical interventions. Posterior distribution of *τ*, the effective start date of NPI, is shown as a histogram of MCMC samples. Red line shows the official last NPI date *τ*∗. Black line shows the estimated 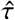. Shaded area shows a 95% credible interval.

We also estimate an early effective start of NPIs in Denmark, Norway, and Sweden. However, the credible intervals are quite wide (Figure 1), and in Denmark and Sweden the evidence did not support the model with free *τ* over the model with *τ* fixed at the official date (WAIC values in Table S1).

In Sweden, the data do not “peak”, i.e., the number of daily cases continues to grow up to Apr 11 (Figure S4a). In Denmark, the opposite occurs: there are seemingly two “peaks”, on Mar 11 and on Apr 8 (Figure S4a); the first “peak” may be a result of stochastic events, for example due to a large cluster of cases or an accumulation of tests. We suspect that these missing and additional “peaks” increase the uncertainty in our inference.

The estimated effective date in Norway is Mar 22, which is two days earlier than the official date of Mar 24. However, the posterior distribution is very wide (±10.79 days 95% CI; Figure 3): it covers the range between Mar 9, three days before the first NPI, and Mar 29, five days after the last NPI (Table 1, Figure S1). The uncertainty in the estimate of the effective start of NPIs might be due to the long duration between the first and last NPIs; however, Germany had the second longest duration between first and last NPIs, but the corresponding posterior distribution is quite narrow (Figure 3).

### Exact effective start of NPIs

We find one case in which the official and effective dates match “*like a Swiss watch*” and the credible interval is narrow. Switzerland ordered a national lockdown on Mar 20, after banning public evens and closing schools on Mar 13 and 14^8^. Indeed, the posterior median 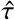 is Mar 19 (±2.51 days 95% CI, see Figure 3). It’s also worth mentioning that Switzerland was the first to mandate self isolation of confirmed cases^8^.

### Assessment of impact of NPIs

The *effective reproduction number R* is the number of secondary cases generated by an infectious case after an epidemic is already underway^4^. We infer the effective reproduction numbers before and after the implementation of NPIs from model parameters (Eq. 7). We then estimate the impact of NPIs as the relative reduction in the reproduction number^8^. We compare the impacts estimated using the fixed *τ* model and the free *τ* model. That is, we compare the impact estimate assuming that NPIs started at the official date *τ*∗, versus the estimate when inferring the effective start of NPIs from the data. Figure 4 demonstrates that estimates from the fixed *τ* model are consistently lower than estimates from the free *τ* model, except in Sweden and Denmark, in which estimation uncertainty is high.

**Figure 4:**
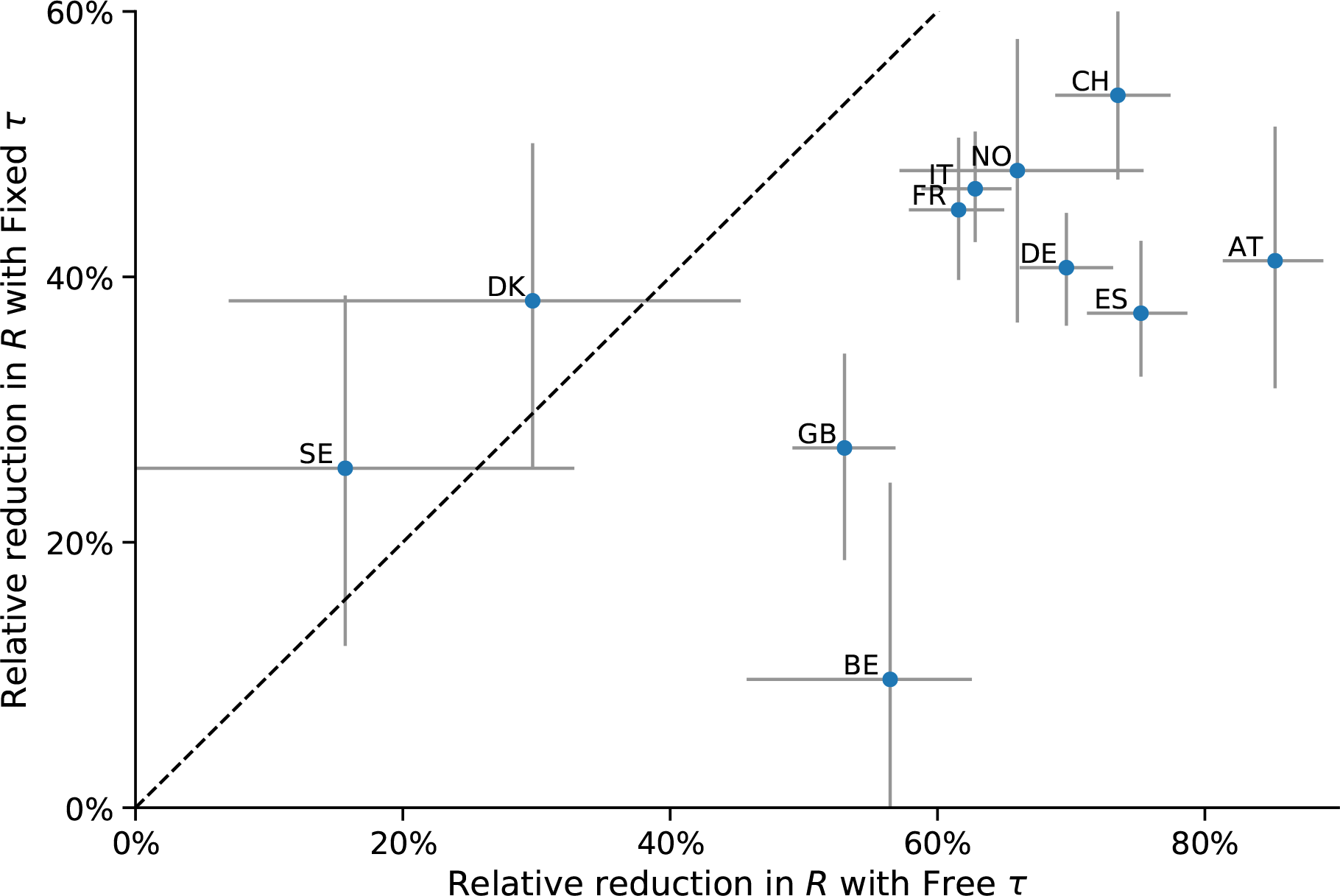
Impact of NPIs is under-estimated when assuming they start at the official date. Shown are estimates of the relative reductions in *R* (the effective reproduction number), which measures the impact of NPIs on disease transmission. The y-axis shows estimates when assuming the start of NPIs is fixed at the official date (fixed *τ*); the x-axis shows estimates when inferring the effective start of NPIs from the data (free *τ*). The dashed line shows a one-to-one correspondence. Markers and bars denote the posterior median and 50% credible intervals. The relative reductions in *R* are consistently lower for the fixed *τ* model (below the dashed line), except in Sweden and Denmark in which uncertainty is high.

These results suggest that the impact of past NPIs by health officials and researchers is likely to be under-estimated if they assume NPIs effectively start at their official dates. The estimated impact can then be interpreted as ineffectiveness of the NPIs, leading to further escalations.

Health officials might also assess the impact of NPIs by comparing model expectations to actual number of daily cases at a specific number of days after the intervention began. However, a significant difference between the official and effective start of interventions can invalidate such assessments. This is illustrated in Figure 5 using data and parameters from Italy: a lockdown was officially ordered on Mar 10 (*τ*∗), but its late effect on the infection dynamics starts on Mar 17 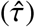. If health officials assume that the dynamics change exactly at *τ*∗, they would expect the number of cases to be within the red lines (posterior predictions assuming *τ* = *τ*∗). This would lead to a significant under-estimation, which might be interpreted as ineffectiveness of the NPI, leading to further escalations. However, the number of cases would actually follow the blue lines (posterior predictions using 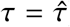), which are a good fit to the real data (stars).

**Figure 5:**
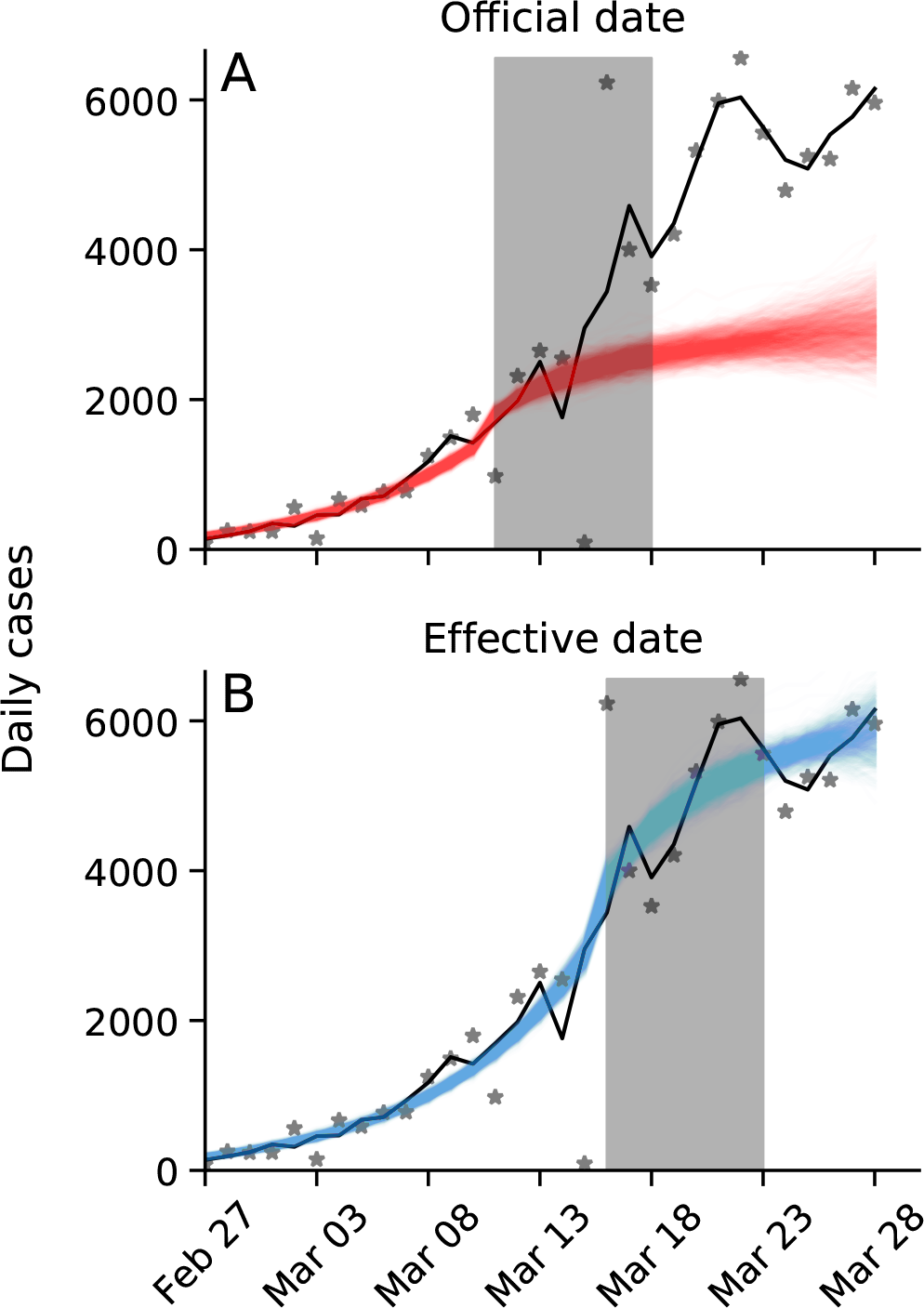
Late effective start of NPIs leads to under-estimation of daily confirmed cases. Real number of daily cases in Italy in black (markers: data; line: time moving average). Model posterior predictions are shown as coloured lines (1,000 draws from the posterior distribution). Shaded box illustrates a generation interval of seven days. **(A)** Using the official date *τ*∗ for the effective start of the NPI, the model under-estimates the number of cases seven days after the start of the NPI. **(B)** Using the estimated date 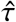 for the effective start of the NPI, the model precisely estimates the number of cases seven days after the start of the NPI. Here, model parameters are best estimates for Italy (Table 2).

## Discussion

We have inferred the effective start date of NPIs in several geographical regions using an SEIR model under an MCMC parameter estimation framework. We find examples of both late and early effective start of NPIs relative to the official date (Figure 1).

In most investigated countries we find a late effective start of NPIs. For example, in Italy and in Wuhan, China, the effective start of the lockdowns seems to have occurred five or more days after the official date (Figure 2). This difference might be explained, in some cases, by low compliance: In Italy, for example, the government plan to implement a lockdown in the Northern provinces leaked to the public, resulting in people leaving these provinces before the lockdown started^10^. Late effect of NPIs may also be due to the time required by both the government and the citizens to prepare for a lockdown, and for new guidelines to be adopted by the population.

In contrast, in some regions we inferred reduced transmission rates even before official lockdowns were implemented, although this is only conclusive in Germany (Figure 3). An early effective date might be due to early adoption of social distancing and similar behavioural adaptations in parts of the population. Adoption of these behaviours may occur via media and social networks, rather than official government recommendations and instructions, and may be influenced by increased risk perception due to domestic or international COVID-19-related reports^1^. Indeed, the evidence supports a change in infection dynamics (i.e. a model with fixed or free *τ*) even for Sweden (Table S1, Table S2, Figure S4a), where a lockdown was not implemented∗.

As expected, we have found that the evidence supports a model in which the transmission rate changes at a specific time point over a model with a constant transmission rate (Table S1 and Table S2). It may be interesting to check if the evidence favors a model with *two* or more change-points, rather than one. Multiple change-points could reflect escalating NPIs (e.g. school closures followed by lockdowns), or an intervention followed by a relaxation. However, interpretation of such models will be harder, as multiple change-points are also likely to result in parameter unidentifiability, for example due to simultaneous implementations of NPIs^8^.

As different countries experiment with various intervention strategies, we expect similar shifts to occur: in some cases the population will be late to comply with new guidelines, whereas in other cases the population will adopt either restrictions or relaxations even before they are formally announced. Attempts to asses the impact of NPIs^3,8^ generally assume they start at their official date, and that a significant change in the dynamics can be observed roughly seven days after the start of NPIs (due to the characteristic generation interval of COVID-19^4^). However, late and early effective start of NPIs, such as we have inferred, can bias these assessments and lead to wrong conclusions about the impact of NPIs (Figure 4, Figure 5).

Our results highlight the complex interaction between personal, regional, and global determinants of behavioral response to an epidemic. Therefore, we emphasize the need to further study variability in compliance and behavior over both time and space. This can be accomplished both by surveying differences in compliance within and between populations^2^, and by incorporating specific behavioral models into epidemiological models^1,6,19^.

## Models and Methods

### Data

We use daily confirmed case data **X** = (*X*_1_, …, *X*_*T*_) from 12 regions during Jan–Apr 2020. These incidence data summarise the number of individuals *X*_*t*_ tested positive for SARS-CoV-2 (using RT-qPCR tests) on each day *t*. Data for Wuhan, China retrieved from Pei and Shaman^16^. Data for 11 European countries retrieved from Flaxman et al.^8^. Where there were multiple sequences of days with zero confirmed cases (e.g. France), we cropped the data to begin with the last sequence so that our analysis focuses on the first sustained outbreak rather than isolated imported cases. For official NPI dates see Table 1.

### SEIR model

We model SARS-CoV-2 infection dynamics by following the number of susceptible *S*, exposed *E*, reported infected *I*_*r*_, unreported infected *I*_*u*_, and recovered *R* individuals in a population of size *N*. This model distinguishes between reported and unreported infected individuals: the reported infected are those that have enough symptoms to eventually be tested and thus appear in daily case reports, to which we fit the model. This model is inspired by Li et al.^15^ and Pei and Shaman^16^, who used a similar model with multiple regions and constant transmission and reporting rates to study COVID-19 dynamics in China and in the continental US.

Susceptible (*S*) individuals become exposed due to contact with reported or unreported infected individuals (*I*_*r*_ or *I*_*u*_) at a rate *β*_*t*_ or *µβ*_*t*_, respectively. The parameter 0 < *µ* < 1 represents the decreased transmission rate from unreported infected individuals, who are often subclinical or even asymptomatic^7,18^. The transmission rate β_*t*_≥ 0 may change over time *t* due to behavioural changes of both susceptible and infected individuals. Exposed individuals, after an average latent period of *Z* days, become reported infected with probability α_*t*_ or unreported infected with probability (1 − *α*_*t*_). The reporting rate 0 < *α*_*t*_ < 1 may also change over time due to changes in human behaviour. Infected individuals remain infectious for an average period of *D* days, after which they either recover, or become ill enough to be quarantined. In either case, they no longer infect other individuals, and therefore effectively become recovered (*R*). The model is described by the following set of equations:

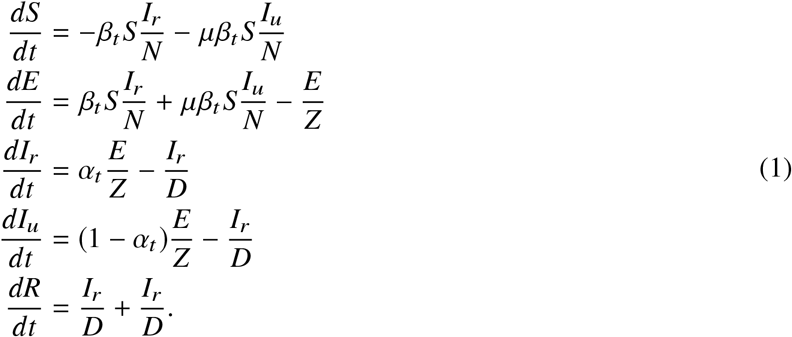

The initial numbers of exposed *E*(0) and unreported infected *I*_*u*_(0) are free model parameters (i.e. inferred from the data), whereas the initial number of reported infected and recovered is assumed to be zero, *I*_*r*_ (0) = *R*(0) = 0, and the number of susceptible is *S*(0) = *N* − *E*(0) − *I*_*u*_(0).

### Likelihood function

For a given vector *θ* of model parameters the *expected* cumulative number of reported infected individuals (*I*_*r*_) until day *t*, following Eq. 1, is

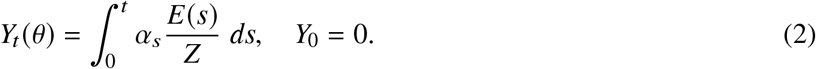

We assume that reported infected individuals are confirmed and therefore observed in the daily case report of day *t* with probability *p*_*t*_ (note that an individual can only be observed once, and that *p*_*t*_ may change over time, but *t* is a specific date rather than the time elapsed since the individual was infected). We denote by *X*_*t*_ the *observed* number of confirmed cases in day *t*, and by 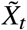 the cumulative number of confirmed cases until end of day *t*,

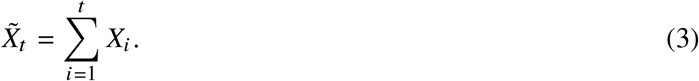

Therefore, at day *t* the number of reported infected yet-to-be confirmed individuals is 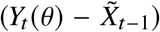. We therefore assume that *X*_*t*_ conditioned on 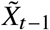 is Poisson distributed, such that

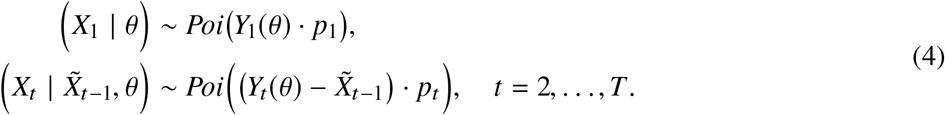

Hence, the *likelihood function* ℒ θ |**X** for a parameter vector θ given the confirmed case data **X** = *X*_1_, …, *X*_*T*_ is defined by the probability to observe **X** given θ,

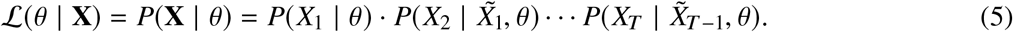

### NPI model

To model non-pharmaceutical interventions (NPIs), we set the start of the NPIs to day *τ* and define

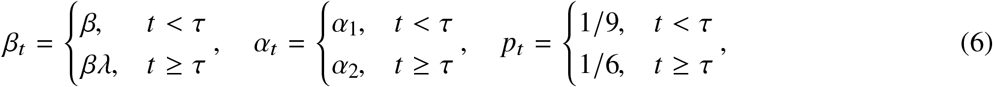

where 0 < *λ* < 1. The values for *p*_*t*_ follow Li et al.^15^, who estimated the average time between infection and reporting in Wuhan, China, at 9 days before the start of NPIs and 6 days after start of NPIs.

Following Li et al.^15^, the effective reproduction numbers before and after the start of NPIs are

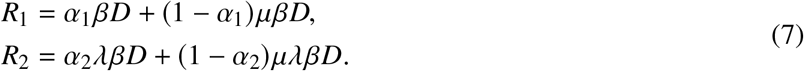

The relative reduction in the effective reproduction number due to NPIs is 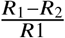.

### Parameter estimation

To estimate the model parameters from the daily case data **X**, we apply a Bayesian inference approach. We start our model Δ*t* days^10^ before the outbreak (defined as consecutive days with increasing confirmed cases) in each country. The model in Eq. 1 is parameterised by the vector θ, where

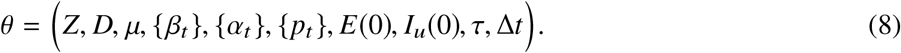

The likelihood function is defined in Eq. 5. The posterior distribution of the model parameters *P* θ **X** is estimated using the affine-invariant ensemble sampler for Markov chain Monte Carlo (MCMC)^12^ implemented in the emcee Python package^9^.

We defined the following prior distributions on the model parameters *P*(*θ*):

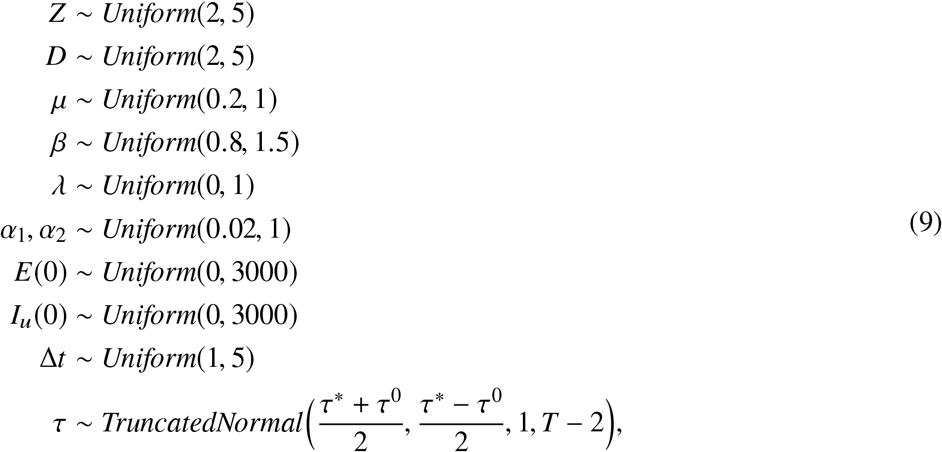

where the prior for *τ* is a truncated normal distribution shaped so that the date of the first and last NPI, *τ*^0^ and *τ*∗ (Table 1), are at minus and plus one standard deviation, and taking values only between 1 and *T* − 2, where *T* is the number of days in the data **X**. We also tested an uninformative uniform prior, *Uniform (*1, *T* − 2). WAIC (Eq. 10) of a model with this uniform prior was either higher, or lower by less than 2, compared to WAIC of a model with the truncated normal prior. The uninformative prior resulted in non-negligible posterior probability for unreasonable *τ* values, such as Mar 1 in the United Kingdom. This was probably due to MCMC chains being stuck in low posterior regions of the parameter space. We therefore decided to use the more informative truncated normal prior for *τ*. Other priors follow Li et al.^15^, with the following exceptions. λ is used to ensure transmission rates are lower after the start of the NPIs (*λ* < 1). We checked values of Δ*t* larger than five days and found they generally produce lower likelihood and unreasonable parameter estimates, and therefore chose *Uniform* 1, 5 as the prior for Δ*t*. We also tried to estimate the value of *p*_*t*_ before and after *τ*, instead of keeping it fixed at 1 9 and 1 6. The model with fixed values was supported by the evidence (lower WAIC, see below) in 9 of 12 countries. Moreover, the estimates for Wuhan, China were 1 9 and 1 6, as in Li et al.^15^. Model fitting was calibrated for case data up to Mar 28, and then applied to data up to Apr 11.

### Model comparison

We perform model selection using two methods. First, we compute WAIC (widely applicable information criterion)^11^,

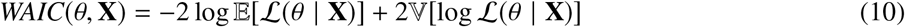

where 𝔼 [·] and 𝕍 [·] are the expectation and variance operators taken over the posterior distribution *P* θ **X**. We compare models by reporting their relative WAIC; lower is better (Table S1). A minority (<5%) of MCMC chains that fail to fully converge can lead to overestimation of the variance (the second termin Eq. 10). Therefore, we exclude from the computation of WAIC chains with mean log-likelihood that is three standard deviations or more from the overall mean.

We also plot posterior predictions: we sample 1,000 parameter vectors from the posterior distribution *P* θ **X** fitted to data up to Apr 11, use these parameter vectors to simulate the SEIR model (Eq. 1) up to Apr 24, and plot the predicted dynamics (Figure S4a). Both the accuracy (i.e. overlap of data and prediction) and the precision (i.e. the tightness of the predictions) are good ways to visually compare models. We also compute the expected posterior RMSE (root mean squared error) of these predictions (Table S2).

### Source code

We use Python 3 with the NumPy, Matplotlib, SciPy, Pandas, Seaborn, and emcee packages. All source code will be publicly available under a permissive open-source license at github.com/yoavram-lab/EffectiveNPI.

## Data Availability

Using publicly available data.

http://github.com/yoavram-lab/EffectiveNPI

## Acknowledgements

We thank Yinon M. Bar-On, Lilach Hadany, and Oren Kolodny for discussions and comments. This work was supported in part by the Israel Science Foundation (3811/19, 552/19, and 1399/17).

## Supplementary Material

**Figure S1:**
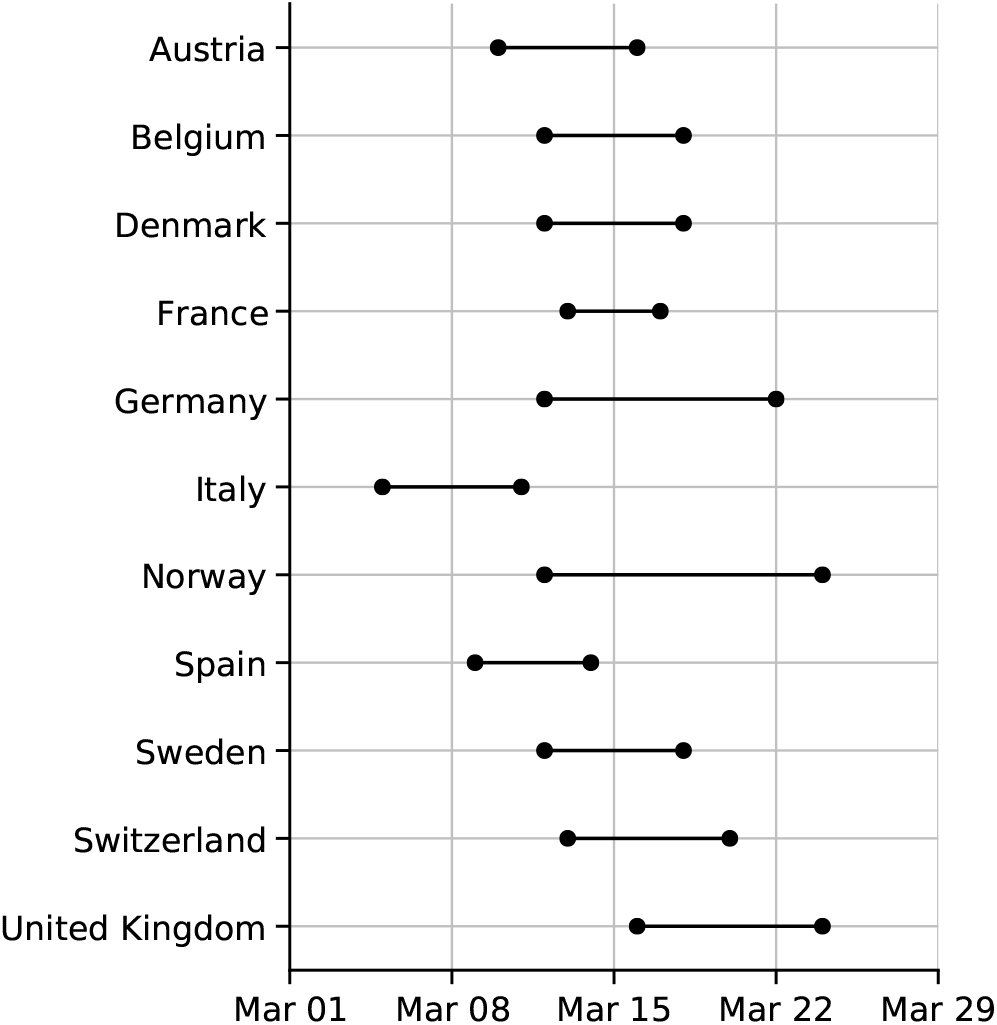
Official start of non-pharmaceutical interventions. See Table 1 for more details. Wuhan, China is not shown.

**Figure S2:**
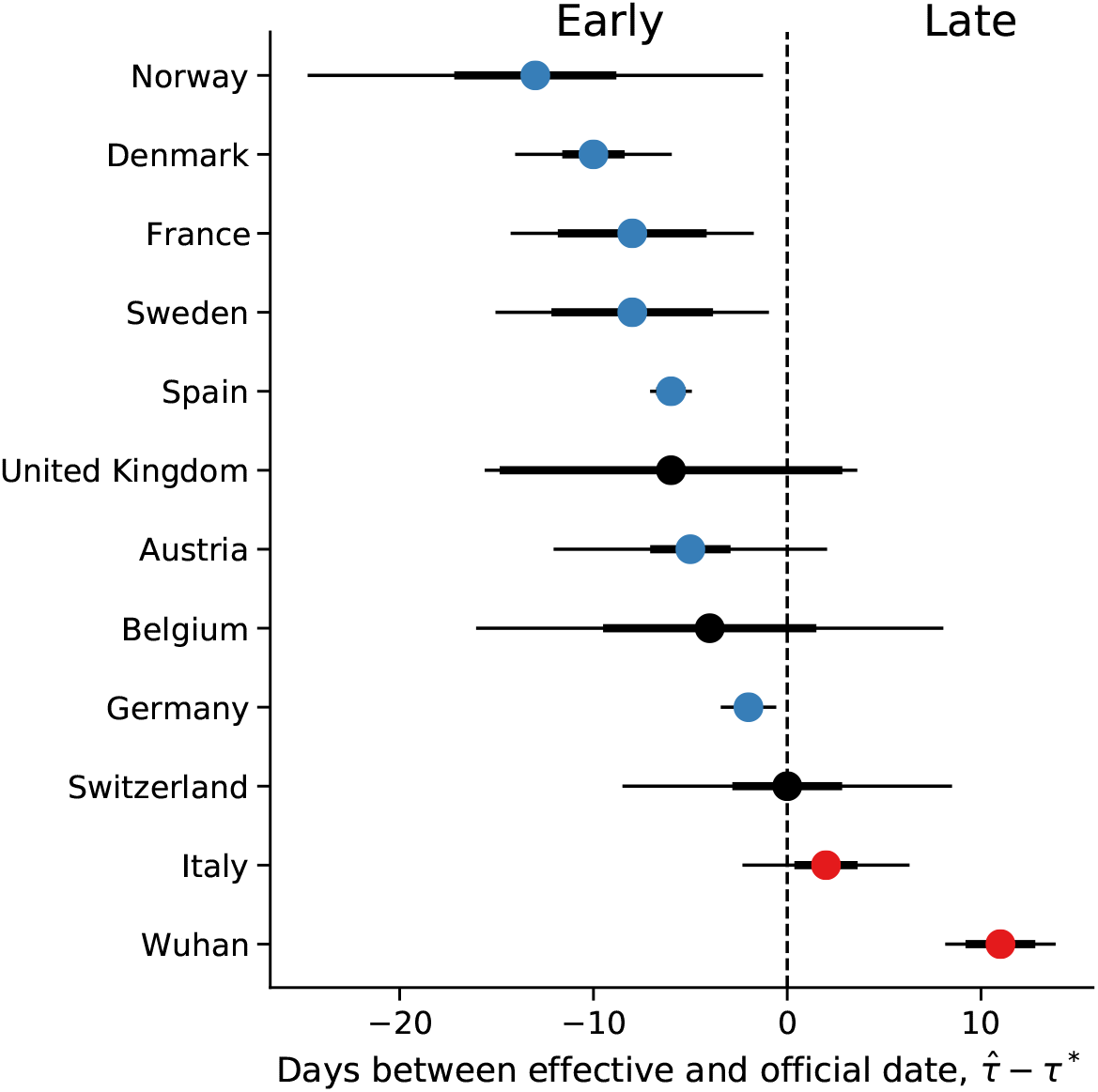
Official vs. effective start of non-pharmaceutical interventions estimated up to Mar 28. The difference between 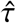 the effective and *τ*∗ the official start of NPIs estimated from case data up to Mar 28, 2020, shown for different regions. Here, 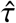 is the posterior median, see Table 2. *τ*∗ is the last NPI date (Table 1). Thin and bold lines show 95% and 75% credible intervals, respectively.

**Table S1:** WAIC values for the different models. WAIC (widely applicable information criterion; Eq. 10)^11^ values for models with: no *τ* at all, *No*; *τ* fixed at the official last NPI date *τ*∗, *Fixed*; and free parameter *τ, Free*. WAIC values are scaled as a deviance measure: lower values imply higher predictive accuracy and a difference of 2 is a popular threshold for model comparison^13^. Bold values emphasize cases in which the *Free* model has the lowest WAIC.

| Country | No | Fixed | Free |
| --- | --- | --- | --- |
| Austria | 219.49 | 95.06 | **35.96** |
| Belgium | 148.37 | 98.41 | **49.16** |
| Denmark | 44.36 | **40.30** | 43.11 |
| France | 581.59 | 255.14 | **172.08** |
| Germany | 1029.36 | 327.50 | **174.90** |
| Italy | 898452.34 | 5484.56 | **80.18** |
| Norway | 70.03 | 42.04 | **39.79** |
| Spain | 1476.46 | 647.34 | **128.58** |
| Sweden | 32.53 | **30.06** | 31.10 |
| Switzerland | 265.80 | 83.95 | **63.89** |
| United Kingdom | 258.18 | 117.54 | **68.17** |
| Wuhan China | 107.31 | 94.00 | **73.75** |

**Table S2:** Posterior RMSE of out-of-sample predictions with the different models. Expected posterior predictive RMSE (root mean squared error) for models with: no *τ* at all, *No*; *τ* fixed at the official last NPI date *τ*∗, *Fixed*; and free parameter *τ, Free*. In all cases, the model with free parameter *τ* has the lowest RMSE. Models were fitted to case data up to Apr 11, 2020, and then used to generate 1,000 predictions up to Apr 24 by sampling model parameters from the posterior distribution. These predictions were then compared to the real data using RMSE, and the mean RMSE value is shown in the table for each country and model. Bold values emphasize cases in which the *Free* model has the lowest RMSE.

| Country | No | Fixed | Free |
| --- | --- | --- | --- |
| Austria | 926.7 | 740.7 | **58.9** |
| Belgium | 3286.0 | 2541.0 | **902.9** |
| Denmark | 434.8 | 1058.0 | **400.0** |
| France | 10530.0 | 8206.0 | **1816.0** |
| Germany | 14240.0 | 17760.0 | **1952.0** |
| Italy | 14160.0 | 9536.0 | **1106.0** |
| Norway | 292.2 | 578.2 | **63.3** |
| Spain | 17840.0 | 14410.0 | **1704.0** |
| Sweden | 775.9 | 1205.0 | **596.4** |
| Switzerland | 1839.0 | 1721.0 | **214.0** |
| United Kingdom | 14700.0 | 14780.0 | **2735.0** |

**Figure S3:**
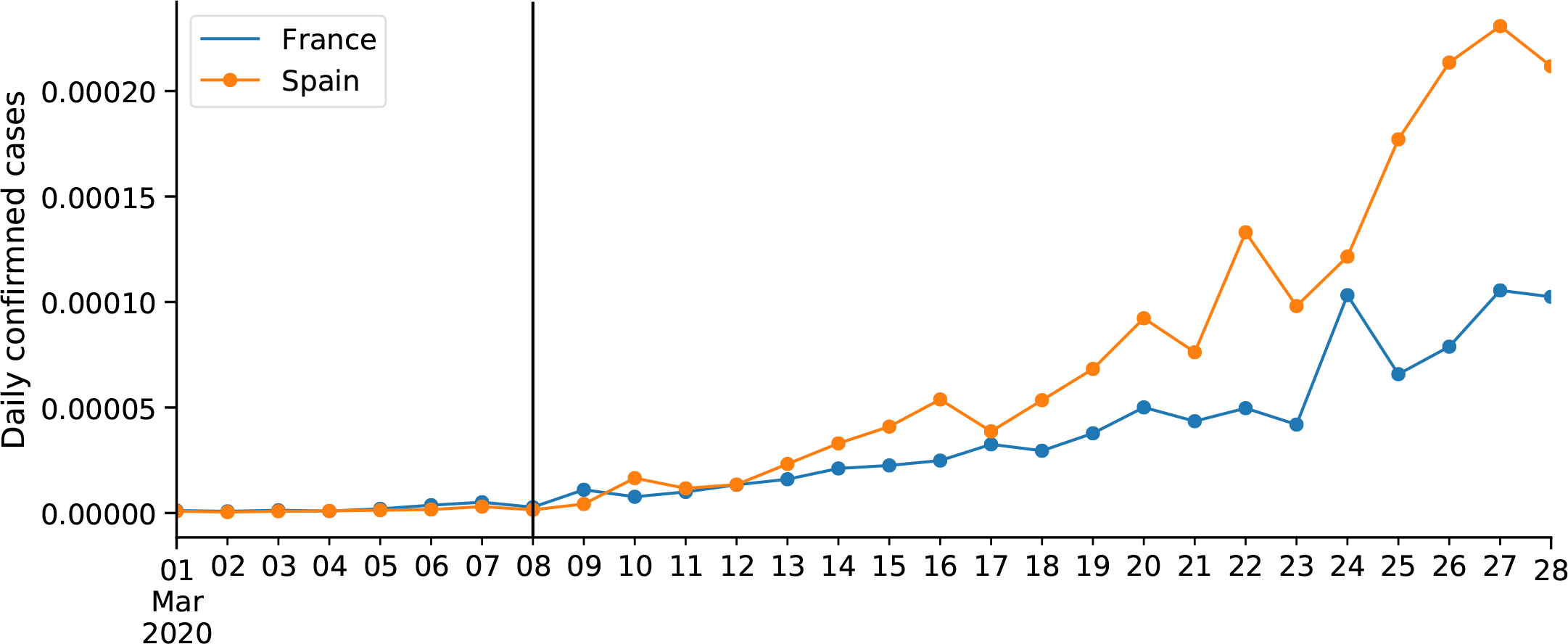
COVID-19 confirmed cases in France and Spain. Number of cases proportional to population size (as of 2018). Vertical line shows Mar 8, the effective start of NPIs 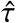 in both countries.

**Figure S4.**
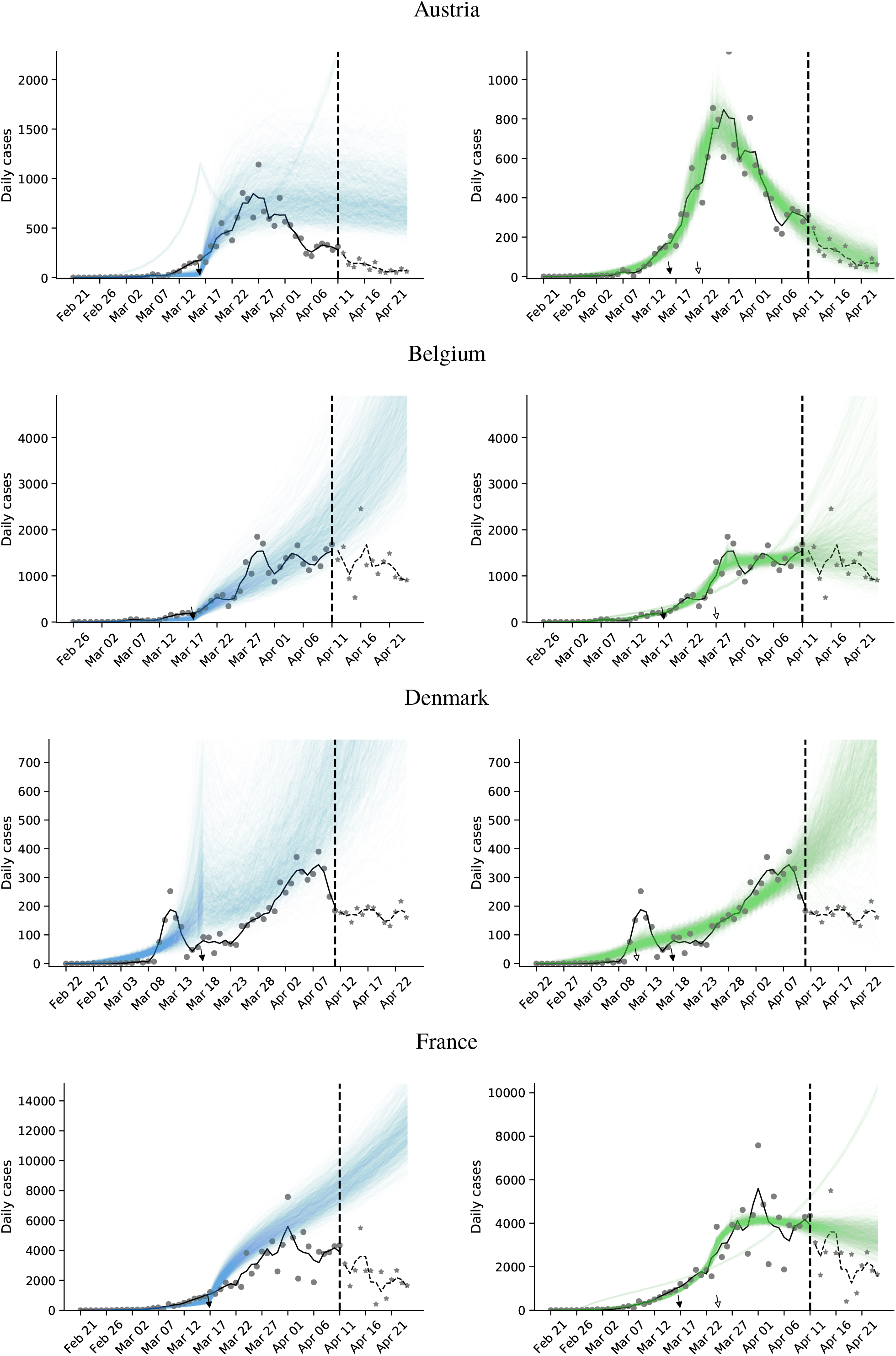
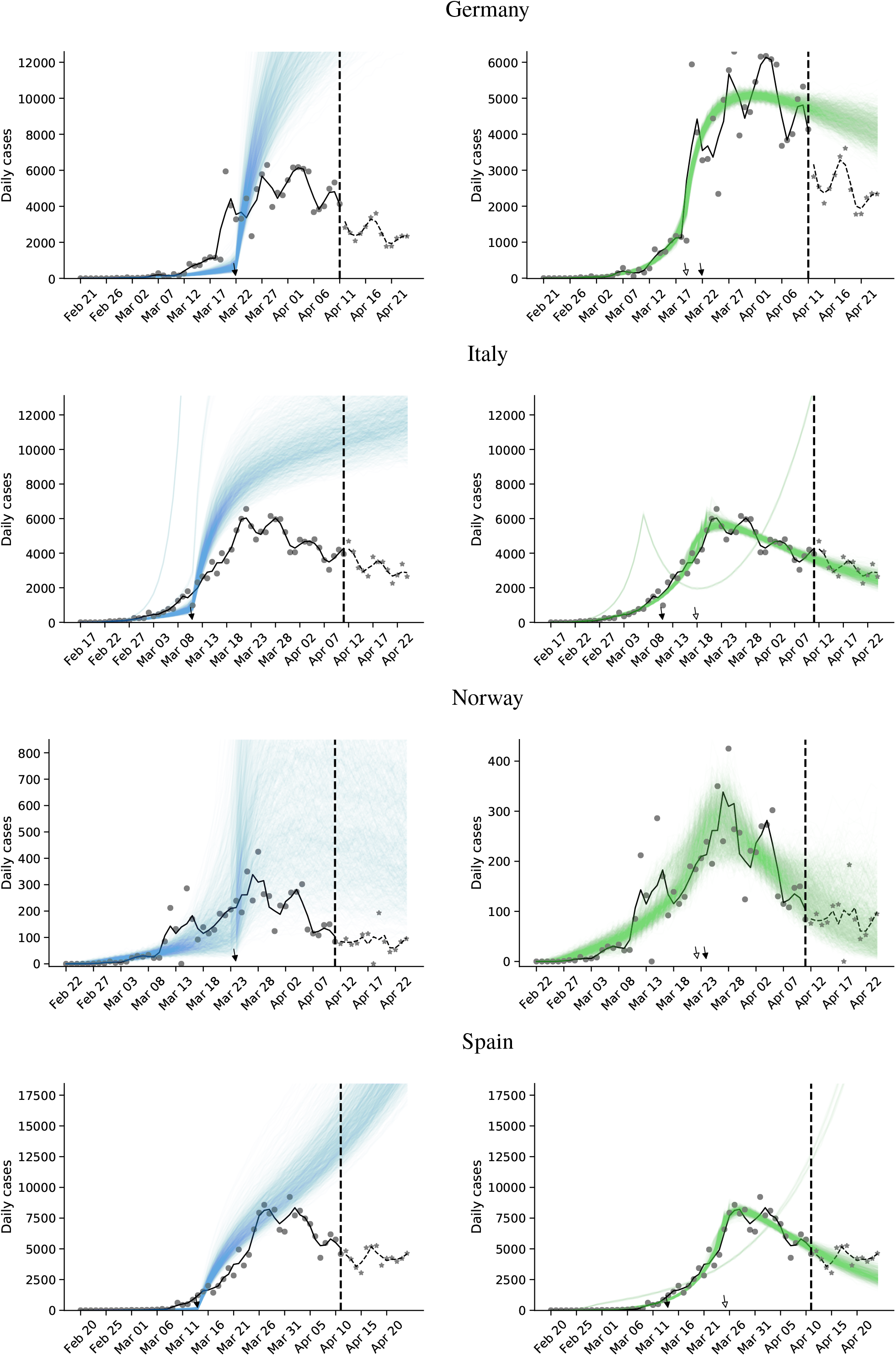
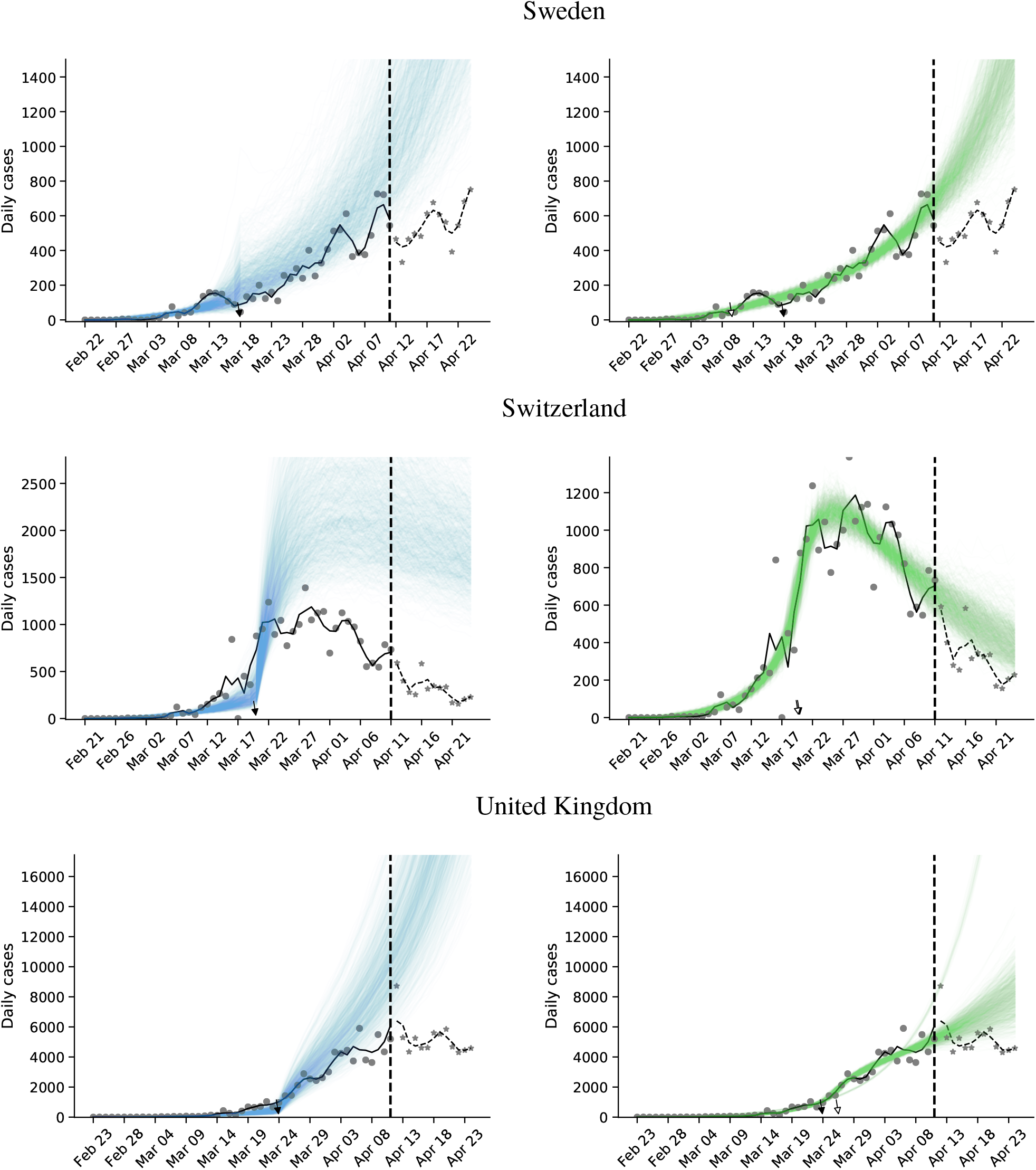
Posterior prediction plots for 11 European countries. The vertical dashed line represents Apr 11, 2020. Circles and stars represent daily case data up to and after Apr 11, respectively Black and white arrows denote the official *τ*∗ and effective 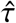 start of NPIs, respectively. Black lines represent a smoothing of the data points using a Savitzky-Golay filter. Coloured lines represent posterior predictions from a model with fixed *τ* (blue) and free *τ* (green). Models were fitted with data up to Apr 11. The predictions are generated by drawing 1,000 parameters sets from the posterior distribution, and then generating a daily case count using the SEIR model up to Apr 24. Note the differences in the y-axis scale. Posterior predictions with the free *τ* model predict the out-of-sample data well for all countries except Denmark and Sweden, but poorly for the fixed *τ* model. The predictions of the model without *τ* (not shown) are even worse.

## Footnotes

∗ Sweden banned public events on Mar 12, encouraged social distancing on Mar 16, and closed schools on Mar 18^8^.

